# Change of prognostic nutritional index after three months of initial hemodialysis is a predictor of long-term outcomes for patients with uremia

**DOI:** 10.64898/2026.08.05.26359770

**Authors:** Shenghui Fu, Haomiao Zhang, Haifei Xie, Fenxia Wang, Lu Bai, Feifei Zhao, Liu Yang, Qing Zhang, Mingfan Lv, Yang Xue, Xiaoluan Liu, Shan Gao, Xinxin Zhang, Pengcheng Xu, Junya Jia

## Abstract

Nutritional status and immune function have a significant impact on the prognosis of patients undergoing maintenance hemodialysis (MHD). Previous studies have shown that the Geriatric Nutritional Risk Index (GNRI) and the Prognostic Nutritional Index (PNI) at the initiation of dialysis can be used to assess the prognosis of MHD. However, as the physical status of patients are usually unstable in the early stage of dialysis, we hypothesized that the nutritional status and immune function after a certain period of stable dialysis might be more closely related to the prognosis. This study conducted a retrospective analysis of patients who started MHD between January 1, 2019 and December 31, 2021. A total of 200 patients were included, with 66 patients succumbing during follow-up. Both initial PNI and initial GNRI exhibited a negative correlation with all-cause mortality (p=0.019 and p=0.046, respectively). After three months of MHD, both PNI and GNRI increased in most patients; however, only the PNI measured after three months was significantly associated with prognosis and higher PNI was associated with a better prognosis (p<0.001). Multivariate Cox regression analyses indicated that only PNI after three months of MHD was linked to prognosis (p=0.004). Kaplan-Meier curves demonstrated patients experiencing a decrease in PNI following three months of MHD had poorer prognoses compared to those whose PNI increased (p=0.004). Furthermore, the predictive value of PNI after three months of MHD was evident in both younger (<60 years old; p=0.024) and older (>60 years old; p=0.022) patient groups. Both the PNI and GNRI showed a downward trend before death, but only PNI had a significant decline (p=0.03, compared with PNI after three months of MHD). In conclusion, for patients undergoing MHD, the correlation between PNI and prognosis is closer than that of GNRI, and the PNI after three months of MHD is a statistically significant but moderate predictor of long-term outcomes.

## Introduction

Maintenance hemodialysis (MHD) is crucial for extending the lives of patients with end-stage renal disease (ESRD) [1, 2]. However, the life expectancy of individuals undergoing hemodialysis is significantly shorter than that of the general population, influenced by various factors that may affect prognosis [3–6]. Researches have demonstrated a correlation between poor nutritional status and unfavorable outcomes in hemodialysis patients. The Geriatric Nutritional Risk Index (GNRI) is an index derived from parameters such as serum albumin concentration, height, and weight. It is primarily utilized to evaluate the nutritional status of elderly individuals and to predict mortality risk across different diseases [7, 8]. In recent years, the GNRI has also been employed to assess prognostic outcomes in MHD [9–11]. Nevertheless, since the GNRI was originally developed for the elderly population, its direct application to all hemodialysis patients warrants caution. Additionally, it is important to recognize that besides nutritional status, multiple factors also contribute to dialysis outcomes.

The Prognostic Nutritional Index (PNI) is an index derived from serum albumin levels and total peripheral lymphocyte counts. Initially developed by the Japanese scholar Onodera, it is also referred to as “Onodera’s index” [12]. The PNI was originally utilized to assess the nutritional and immune status of patients undergoing gastrointestinal surgery; however, it has gradually evolved into a novel indicator for evaluating the prognosis of various diseases, including malignancies of the digestive tract, gynecological tumors, lung cancer, as well as non-malignant issues such as fractures, heart failure, and cerebral infarction [13]. Several studies have investigated the use of PNI for predicting outcomes in peritoneal dialysis patients, showing that a low PNI is associated with increased mortality and infection rates in this population [14–19]. However, research on the application of PNI for prognosis assessment in patients undergoing MHD remains relatively limited. Moreover, given that long-term hemodialysis is often accompanied by impaired immune function, PNI may theoretically have an advantage over GNRI in predicting outcomes in this group. The present study therefore aims to compare the prognostic value of PNI and GNRI in the hemodialysis population.

Patients newly initiated on MHD typically undergo a period of physiological adaptation and are susceptible to various dialysis-related complications. Some may experience early mortality due to severe underlying diseases or other complex factors. Therefore, most clinical studies involving MHD patients require a stable dialysis duration of more than three months. In this study, we specifically calculated the GNRI and PNI values at both baseline and three months after dialysis initiation, and subsequently compared the predictive value of these four indicators for the endpoint of all-cause mortality in MHD patients.

## Materials and methods

### Patients

Patients who commenced MHD at the Hemodialysis Center of Tianjin Medical University General Hospital and who underwent systematic laboratory evaluations in our center prior to initiating dialysis were included in this study. All participants were required to have received stable hemodialysis for a minimum duration of three months. A retrospective analysis was performed in our hospital from January 1st, 2019, to December 31, 2021. We accessed the data for research purposes on December 1st, 2024. The follow-up period concluded on June 30, 2024. We had access to information that could identify individual participants during or after data collection.

Patients with comorbidities such as malignant tumors, active infections, or autoimmune diseases that significantly impact survival were excluded from the study. Additionally, those who did not undergo timely re-examination after three months of dialysis were also excluded. This research was conducted in accordance with the ethical principles outlined in the Helsinki Declaration and received approval from the Research Ethics Committee of Tianjin Medical University General Hospital (approval code: IRB2024-YX-571-01; date: November 28, 2024). The requirement for informed consent was waived due to the retrospective nature of the study and the use of anonymized data.

### Data Collection

The following clinical and laboratory parameters were collected: age, gender, mean arterial pressure, history of cardiovascular diseases, history of diabetes mellitus, initial serum creatinine levels, serum white blood cell count, serum neutrophil count, serum lymphocyte count (both initial data and data after three months of dialysis), serum platelet count, serum hemoglobin concentration, serum albumin levels (initial data and data after three months of dialysis), serum globulin levels, as well as measurements for serum potassium, sodium, phosphorus, calcium, total cholesterol, total triglycerides, high-density lipoprotein (HDL), low-density lipoprotein (LDL), and C-reactive protein (CRP).

Estimated Glomerular filtration rate (eGFR) was calculated by simplified and Modification of Diet in Renal Diseases (MDRD) formula [20].

Body mass index (BMI) was calculated by dividing post-hemodialysis dry weight by baseline height squared (kg/m^2^).

The PNI was calculated using the following formula [21]: PNI=[10×serum albumin (g/dL)] + [0.005×total lymphocyte count (/mm^3^)].

The GNRI was calculated using the following formula [22]: GNRI=[1.489×albumin (g/l)]+41.7×[body weight (kg)/ideal body weight (kg)]; the ideal weight was calculated using the following formula: Ideal body weight (men)=height (cm)×0.75-62.5; and Ideal body weight (women)=height (cm)×0.60-40.

When the body weight exceeded the ideal body weight, the ratio of bodyweight to ideal body weight was set as “1”.

### Statistical analyses

For continuous variables, normally distributed variables were described as means±SDs while obviously skewed variables are expressed as the median (interquartile range, IQR). The proportion or prevalence was used to describe categorical variables. Continuous variables were compared with the independent samples T-test or one-way ANOVA test. Categorical parameters were compared with the χ^2^ test. To evaluate the effect of biomarkers in predicting outcomes, receiver operating characteristic (ROC) curves were plotted with the area under the curve (AUC) and the best cut-off values and Youden’ index were calculated. Multivariate analyses were used to identify independent prognostic variables Parameters that appeared significant in univariate analysis for survival were included in the Cox multivariate regression analysis. The 95% confidence interval (CI) was used to indicate the relationship between survival time and each independent factor. Survival analyses of prognostic indexes, and clinical and pathological features were calculated using the Kaplan–Meier method (log-rank test). Statistical significance level was p<0.05. The Statistical Package for Social Sciences for Windows 21.0 (SPSS, Inc., Chicago, IL, United States) was used for analysis.

## Results

### Patient enrollment

From January 1, 2019, to December 31, 2021, a total of 276 patients commenced MHD at the Hemodialysis Center of Tianjin Medical University General Hospital. Four patients were excluded due to undergoing dialysis for less than three months. Additionally, 37 patients were excluded because they lacked PNI data after three months of dialysis, and another 18 patients were excluded due to missing GNRI data after the same duration. Furthermore, among the remaining 217 patients, 17 individuals were excluded due to comorbidities such as malignant tumors, active infections, or autoimmune diseases that significantly impact life expectancy. Ultimately, a total of 200 patients were included in the study. All patients underwent regular hemodialysis three times a week. Among them, 144 patients used arteriovenous fistulas for dialysis, 53 patients used semi-permanent catheters in the internal jugular vein, and the remaining 3 patients used artificial blood vessels for dialysis. Among these 200 participants, there were 118 males and 82 females. No statistically significant differences were observed between male and female patients regarding age; serum albumin; electrolytes; uric acid levels; total white blood cell count; serum platelet count; serum lymphocyte count; and PNI values. However, compared to females, males exhibited higher levels of hemoglobin (g/L, 85.53±9.86 vs. 80.05±17.05, p=0.044), serum creatinine (μmol/L, 739.89±283.46 vs. 645.16±225.02, p=0.012) and initial eGFR (mL/min/1.73m^2^, 11.28±4.70 vs. 9.26±6.69, p=0.012).

### The relationship between PNI, GNRI and the clinical laboratory parameters

The initial PNI, PNI after 3 months of dialysis, initial GNRI, and GNRI after 3 months of dialysis were calculated. All 200 patients were divided into low initial PNI group and high initial PNI group according to the median of initial PNI. Similarly, all patients were also divided into low initial GNRI group and high initial GNRI group based on the median of initial GNRI. As shown in Table 1, the existence of diabetes had an influence on both initial PNI and initial GNRI. The proportion of patients with diabetes in the low initial PNI group was significantly higher than that in the high initial PNI group (p=0.001), and the proportion of patients with diabetes in the low initial GNRI group was also significantly higher than that in the high initial GNRI group (p=0.009). The existence of coronary heart disease and the blood pressure (expressed by mean arterial pressure) had no influence on either initial PNI or initial GNRI. The serum CRP level also influenced both initial PNI and initial GNRI. The serum CRP level in the low initial PNI group was significantly higher than that in the high initial PNI group (p=0.001), and the serum CRP level in the low initial GNRI group was also significantly higher than that in the high initial GNRI group (p=0.002). It was worth noting that although PNI was calculated based on the counts of blood lymphocytes, there was no difference of the counts of peripheral white blood cells or neutrophils between the low initial PNI group and the high initial PNI group. However, although the calculation of GNRI was not based on the number of any immune cells, the counts of peripheral white blood cells and neutrophils in the low initial GNRI group were significantly higher than those in the high initial GNRI group (p=0.006 and p=0.017). Additionally, it is worth noting that the blood calcium levels of patients in the low initial PNI group were significantly lower than those in the high initial PNI group (p<0.001), while there was no difference of blood calcium levels between patients in the low initial GNRI group and patients in the high initial GNRI group (p=0.474).

**Table 1.** Baseline patient characteristics stratified by median initial PNI and GNRI. BMI: body mass index; CRP: C-reactive protein; GNRI: geriatric nutritional risk index; PNI: prognostic nutritional index.

|  | PNI |  |  | GNRI |  |  |
| --- | --- | --- | --- | --- | --- | --- |
|  | Low initial<br>PNI(n=100) | High initial PNI<br>(n=100) | P value | Low initial<br>GNRI (n=100) | High initial<br>GNRI (n=100) | P value |
| Age(years) | 57.65±15.75 | 52.44±14.81 | 0.051 | 56.39±15.93 | 53.70±14.94 | 0.220 |
| Gender, male(%) | 59% | 59% | 1.000 | 43% | 39% | 0.565 |
| BMI (kg/m<sup>2</sup>) | 23.02±3.70 | 23.20±4.22 | 0.313 | 21.77±3.71 | 24.45±3.75 | 0.001 |
| Mean arterial pressure(mmHg) | 111.24±17.43 | 111.96±16.41 | 0.485 | 110.52±16.68 | 112.68±17.10 | 0.942 |
| Cardiovascular diseases (%) | 28% | 24% | 0.492 | 29% | 23% | 0.312 |
| Diabetes mellitus(%) | 50% | 28% | 0.001 | 48% | 30% | 0.009 |
| Dialysis with catheter (%) | 28% | 25% | 0.631 | 29% | 24% | 0.423 |
| Initial eGFR(mL/min/1.73m<sup>2</sup>) | 11.04±5.31 | 9.85±5.99 | 0.332 | 10.58±5.30 | 10.33±6.04 | 0.866 |
| White blood cells (10<sup>9</sup>/L) | 7.61±4.08 | 7.45±2.60 | 0.737 | 8.19±4.00 | 6.87±2.56 | 0.006 |
| Neutrophils(10<sup>9</sup>/L) | 5.89±3.84 | 5.43±2.07 | 0.287 | 6.178±3.63 | 5.14±2.32 | 0.017 |
| Lymphocytes(10<sup>9</sup>/L) | 0.95±0.48 | 1.25±0.65 | 0.006 | 1.16±0.71 | 1.04±0.43 | 0.168 |
| Platelets(10<sup>9</sup>/L) | 196.63±86.74 | 188.21±98.24 | 0.081 | 197.62±90.03 | 187.22±95.13 | 0.716 |
| Hemoglobin (g/L) | 79.12±20.51 | 87.44±16.21 | 0.182 | 80.28±20.01 | 86.28±17.32 | 0.208 |
| Albumin(g/L) | 26.58±4.65 | 35.57±3.82 | <0.001 | 26.83±4.92 | 35.32±4.06 | <0.001 |
| Globulin(g/L) | 27.63±5.03 | 28.63±4.70 | 0.148 | 27.51±5.20 | 28.75±4.43 | 0.072 |
| Serum potassium(mmol/L) | 4.47±0.87 | 4.61±0.76 | 0.224 | 4.51±0.79 | 4.56±0.84 | 0.308 |
| Serum sodium(mmol/L) | 138.04±4.99 | 139.09±4.08 | 0.287 | 138.03±5.08 | 139.10±3.97 | 0.103 |
| Serum phosphorus (mmol/L) | 1.74±0.51 | 1.83±0.65 | 0.299 | 1.77±0.52 | 1.79±0.64 | 0.435 |
| Serum calcium(mmol/L) | 1.87±0.25 | 2.00±0.26 | <0.001 | 1.92±0.23 | 1.95±0.29 | 0.474 |
| Total cholesterol (mmol/L) | 4.80±0.91 | 4.73±1.23 | 0.893 | 4.80±1.90 | 4.73±1.25 | 0.643 |
| Total triglyceride (mmol/L) | 1.67±0.84 | 1.82±1.01 | 0.305 | 1.66±0.83 | 1.82±1.02 | 0.136 |
| High-density lipoprotein (mmol/L) | 1.11±0.39 | 1.09±0.35 | 0.452 | 1.13±0.41 | 1.06±0.31 | 0.242 |
| Low-density lipoprotein (mmol/L) | 2.86±1.28 | 2.73±0.84 | 0.513 | 2.84±1.23 | 2.74±0.92 | 0.592 |
| Serum CRP(mg/dL) | 0.93(0.37, 4.09) | 0.58(0.19, 1.36) | 0.001 | 0.94(0.35, 3.84) | 0.55(0.19, 1.69) | 0.002 |
| GNRI | 81.43±8.07 | 94.04±8.03 | <0.001 | 79.61±6.48 | 95.86±5.86 | <0.001 |
| PNI | 31.36±4.22 | 41.85±3.41 | <0.001 | 32.65±5.68 | 40.56±4.59 | <0.001 |

### Correlation analysis between GNRI and peripheral lymphocyte counts

Although the calculation of GNRI does not require the parameter of peripheral blood lymphocytes, since we found that GNRI seemed to have some relationship with the peripheral blood white blood cell count when comparing between groups, we analyzed the correlation between the initial GNRI and the GNRI three months after dialysis initiation and the peripheral lymphocytes at the same period. As shown in Fig 1, there was no correlation between the initial GNRI and the initial peripheral lymphocyte count, while the GNRI three months after dialysis initiation had a weak positive correlation with the peripheral lymphocyte count three months after dialysis (p = 0.037).

**Fig 1.**
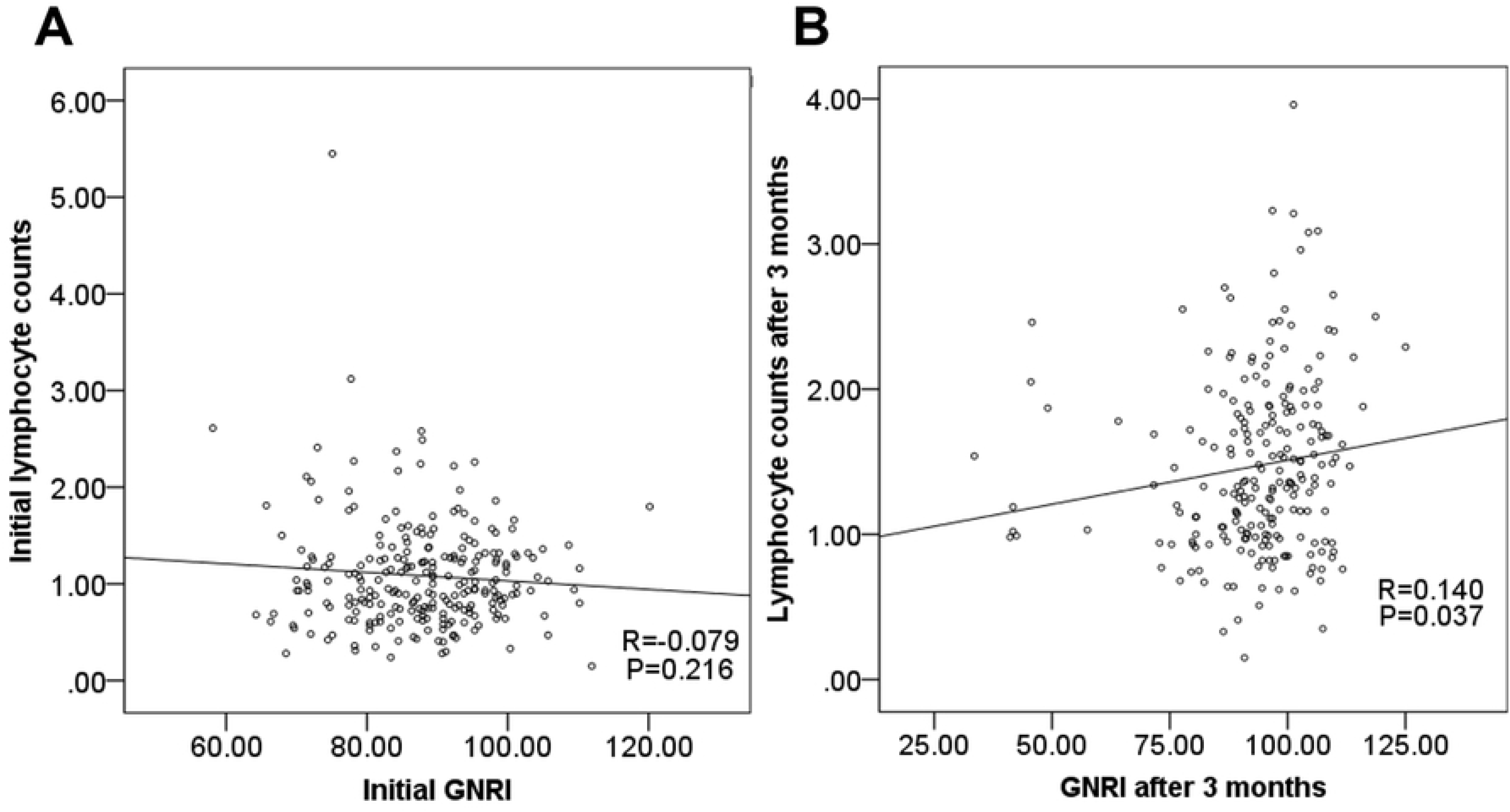
Correlation of initial GNRI (A) and GNRI at three months of dialysis (B) with corresponding peripheral blood lymphocyte counts.

### Relationship between PNI, GNRI and the all-cause mortality

The occurrence of all-cause mortality during the follow-up period was defined as the endpoint event. At the conclusion of the follow-up, 66 patients had died while the remaining individuals survived. Among the 66 deceased patients, 21 succumbed to cardiac diseases, 14 to stroke, 5 to infections, and the causes of death for the remaining 26 were rather ambiguous. To avoid survivor bias, we set the time origin at 3 months after the start of dialysis and performed Cox analysis. As illustrated in Table 2, univariate Cox analyses indicated that both initial PNI and PNI measured after three months of dialysis were significantly associated with all-cause mortality (p=0.019 and p<0.001, respectively). In addition, advanced age, a history of diabetes mellitus, and catheter-based dialysis were also identified as independent risk factors for adverse prognosis. Notably, the association between PNI after three months of dialysis and all-cause mortality was considerably more obvious than that observed for initial PNI. Additionally, the initial GNRI level demonstrated a relationship with all-cause mortality (p=0.046); however, no significant correlation was found between GNRI assessed after three months of dialysis and all-cause mortality (p=0.056). In multivariate Cox analyses, only PNI measured after three months of dialysis remained significantly associated with all-cause mortality. Each 1-point increase of PNI reduced mortality risk by ∼9.2% (HR=0.908). Initial PNI, initial GNRI, and GNRI evaluated after three months of dialysis did not exhibit any further association with prognosis. The variance inflation factor (VIF) for all included variables was calculated to be less than 5, indicating no significant multicollinearity.

**Table 2.** Univariate and multivariate Cox regression analyses of prognostic factors for survival. CRP: C reactive protein; GNRI: geriatric nutritional risk index; PNI: prognostic nutritional index.

|  | Univariate analysis |  |  | Multivariate analysis |  |  |
| --- | --- | --- | --- | --- | --- | --- |
|  | HR | 95% CI | P value | HR | 95% CI | P value |
| Gender (male) | 1.550 | 0.932-2.576 | 0.091 | 1.825 | 1.074-3.099 | 0.026 |
| Age | 1.018 | 1.001-1.034 | 0.036 | 1.007 | 0.990-1.025 | 0.410 |
| Without diabetes | 0.447 | 0.274-0.728 | 0.001 | 0.557 | 0.335-0.926 | 0.024 |
| Dialysis through a catheter | 1.647 | 1.001-2.710 | 0.05 | 0.675 | 0.403-1.133 | 0.137 |
| CRP level | 1.005 | 0.934-1.082 | 0.890 | - | - | - |
| Initial PNI | 0.957 | 0.922-0.993 | 0.019 | 1.034 | 0.967-1.106 | 0.331 |
| PNI after 3 months | 0.938 | 0.905-0.972 | <0.001 | 0.908 | 0.837-0.984 | 0.018 |
| Initial GNRI | 0.977 | 0.954-1.000 | 0.046 | 0.959 | 0.913-1.007 | 0.095 |
| GNRI after 3 months | 0.977 | 0.954-1.001 | 0.056 | 1.045 | 0.987-1.106 | 0.135 |

ROC analysis revealed that the area under the curve (AUC) for initial PNI, PNI after three months of dialysis, initial GNRI, and GNRI after three months of dialysis were 0.635, 0.689, 0.616, and 0.627 respectively (Fig 2). Notably, the AUC for PNI after three months of dialysis was the highest among these metrics. Furthermore, when the initial PNI reached a value of 40.8, the Youden index attained its maximum at 0.351 (Table 3).

**Fig 2.**
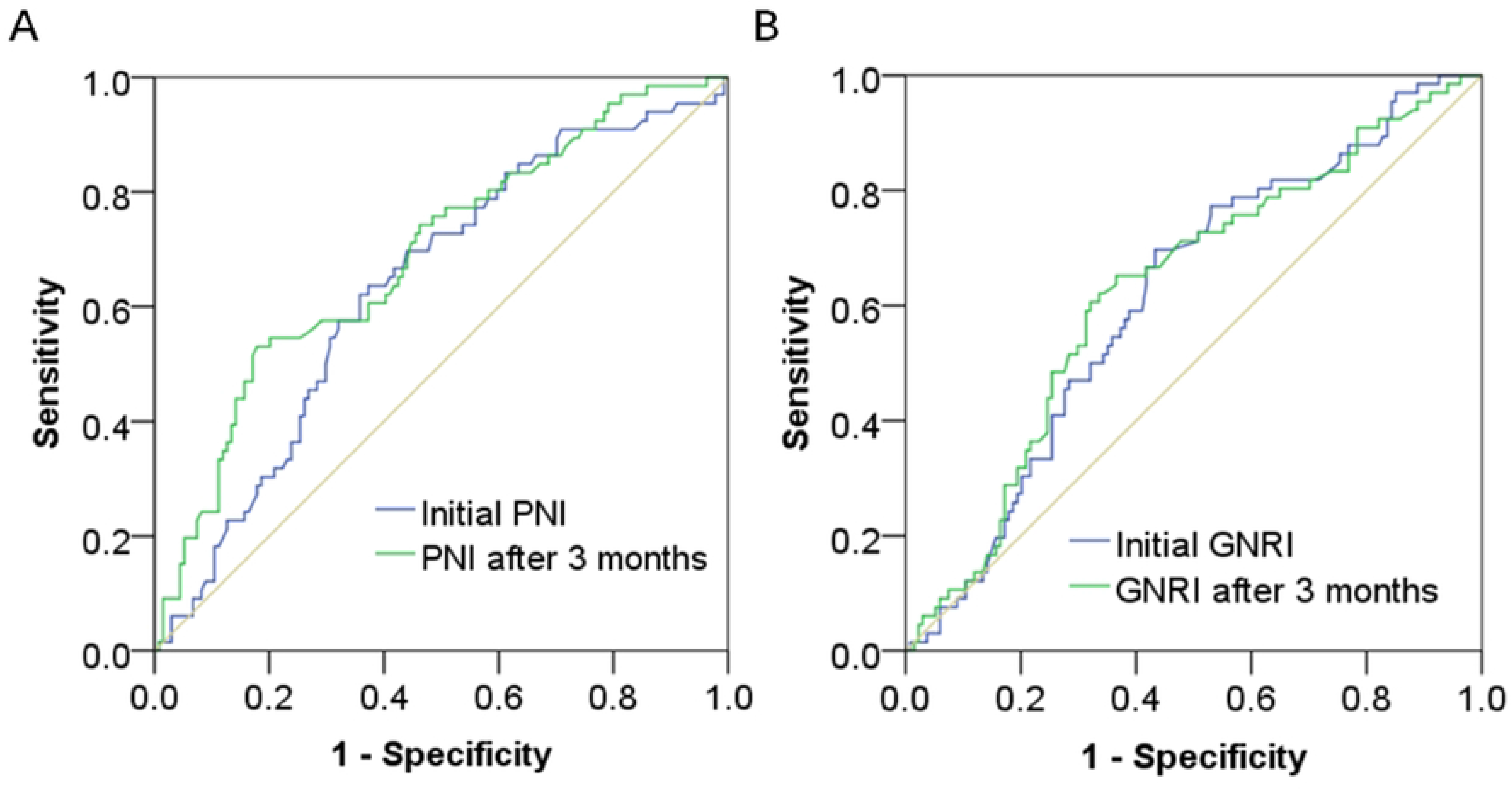
ROC analysis of PNI and GNRI. GNRI: geriatric nutritional risk index; PNI: prognostic nutritional index.

**Table 3.**
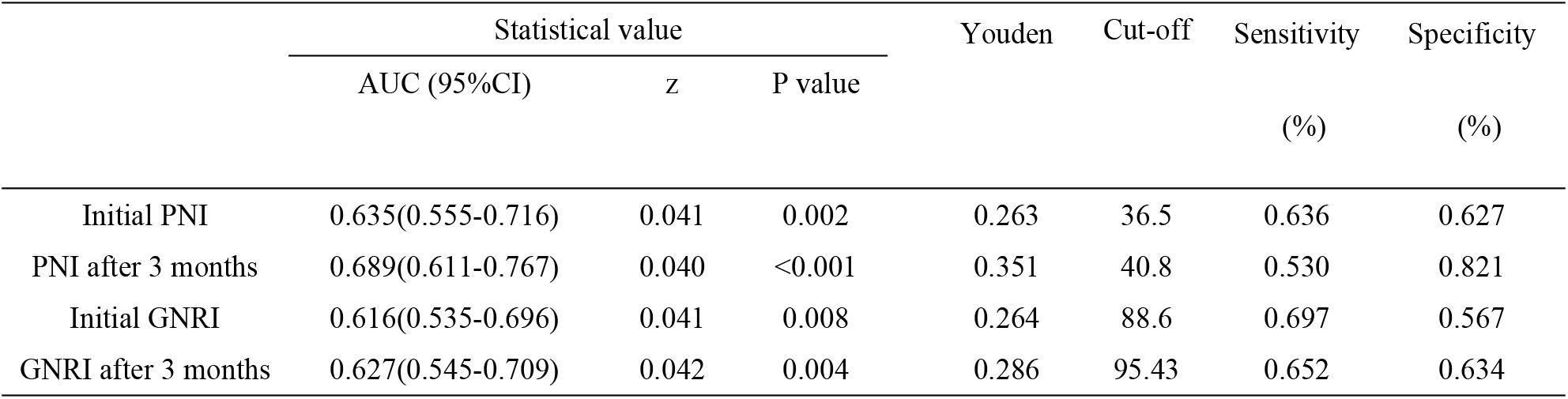
ROC analysis of PNI, PNI at three months, GNRI, and GNRI at three months for predicting all-cause mortality. GNRI: geriatric nutritional risk index; PNI: prognostic nutritional index.

|  | Statistical value |  |  | Youden | Cut-off | Sensitivity | Specificity |
| --- | --- | --- | --- | --- | --- | --- | --- |
|  | AUC (95%CI) | z | P value |  |  | (%) | (%) |
| Initial PNI | 0.635(0.555-0.716) | 0.041 | 0.002 | 0.263 | 36.5 | 0.636 | 0.627 |
| PNI after 3 months | 0.689(0.611-0.767) | 0.040 | <0.001 | 0.351 | 40.8 | 0.530 | 0.821 |
| Initial GNRI | 0.616(0.535-0.696) | 0.041 | 0.008 | 0.264 | 88.6 | 0.697 | 0.567 |
| GNRI after 3 months | 0.627(0.545-0.709) | 0.042 | 0.004 | 0.286 | 95.43 | 0.652 | 0.634 |

### Decreased PNI after 3 months of dialysis was associated with a poor prognosis

According to previous studies, we defined PNI < 38 and GNRI < 98 as the criteria for malnutrition [23]. At the initiation of dialysis, 59% of patients met the criteria for malnutrition according to PNI, while 83% met the criteria according to GNRI. After three months of dialysis, 17% of patients met the criteria for malnutrition according to PNI, whereas 57.5% met the criteria according to GNRI. Among the 200 patients studied, 26 exhibited both a decrease in PNI and a decrease in GNRI after three months of dialysis. Additionally, 3 patients demonstrated an increase in GNRI alongside a decrease in PNI, while 12 patients showed an increase in PNI with a corresponding decrease in GNRI. The remaining 159 patients experienced increases in both PNI and GNRI.

The Kaplan-Meier survival curves demonstrated that after grouping by median PNI and GNRI at dialysis initiation, patients in the high-PNI and high-GNRI groups had better prognoses than those in the low-PNI and low-GNRI groups (Fig 3A-B). Patients exhibiting a decrease in the PNI after three months of dialysis experienced a poorer prognosis compared to those with an increase in PNI. Conversely, when assessing the prognoses of patients with decreased GNRI against those with increased GNRI following three months of dialysis, no statistically significant difference was observed (Fig 3C-D). To eliminate survivor bias, we performed landmark analyses with the time origin set at 3 months post-dialysis, grouping patients by the median PNI and GNRI values measured at that time point, and obtained similar results (Fig 3E-H).

**Fig 3.**
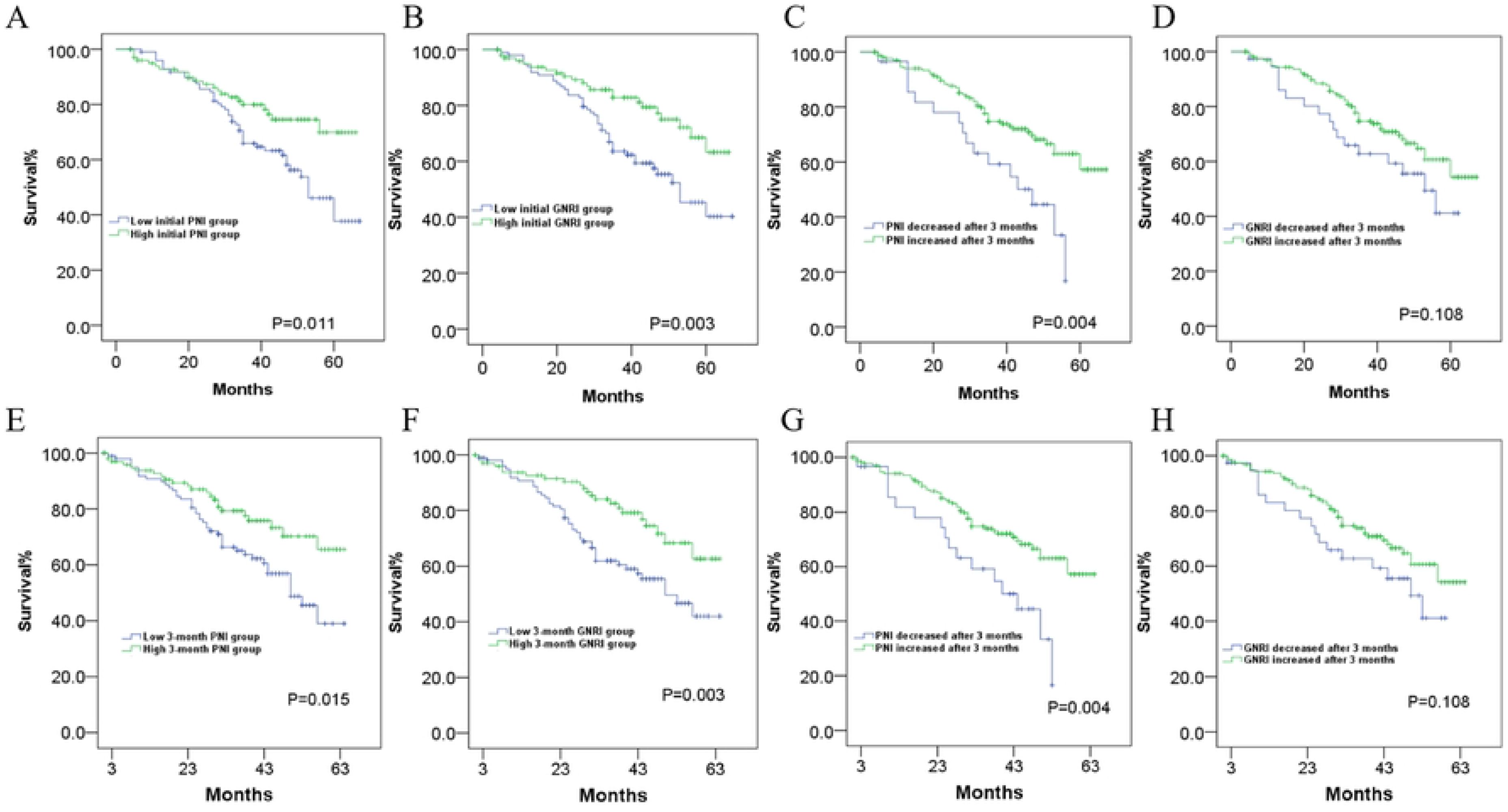
Kaplan-Meier curves for all-cause mortality according to initial PNI (A), initial GNRI (B), PNI at three months of dialysis (C), and GNRI at three months of dialysis (D). E–H: Landmark analyses with time origin at 3 months post-dialysis. E: grouped by median PNI at 3 months; F: grouped by median GNRI at 3 months. GNRI: geriatric nutritional risk index; PNI: prognostic nutritional index.

### Prognostic significance of PNI and GNRI in patients across different age groups

The GNRI was originally developed for elderly patients; therefore, we assessed the predictive value of the PNI and GNRI regarding patient prognosis across different age groups. As illustrated in Fig 4, the initial PNI exhibited a negative correlation with all-cause mortality events among the entire cohort of 200 patients (HR = 0.957, 95% CI: 0.922–0.993, p=0.019). In patients under 60 years of age, the predictive significance of initial PNI was particularly pronounced (HR = 0.920, 95% CI: 0.870–0.973, p=0.004); however, it demonstrated no predictive value in those aged over 60. Furthermore, PNI measured after three months of dialysis showed prognostic relevance for both younger patients under 60 years old (HR = 0.944, 95% CI: 0.898–0.992, p=0.024) and older patients above this age threshold (HR = 0.935, 95% CI: 0.882–0.990, p=0.022). Notably, initial GNRI only displayed predictive value in individuals under the age of sixty; conversely, there was no discernible relationship between initial GNRI and prognosis in those over sixty. Additionally, GNRI assessed after three months of dialysis did not demonstrate any significant predictive capacity for either group.

**Fig 4.**
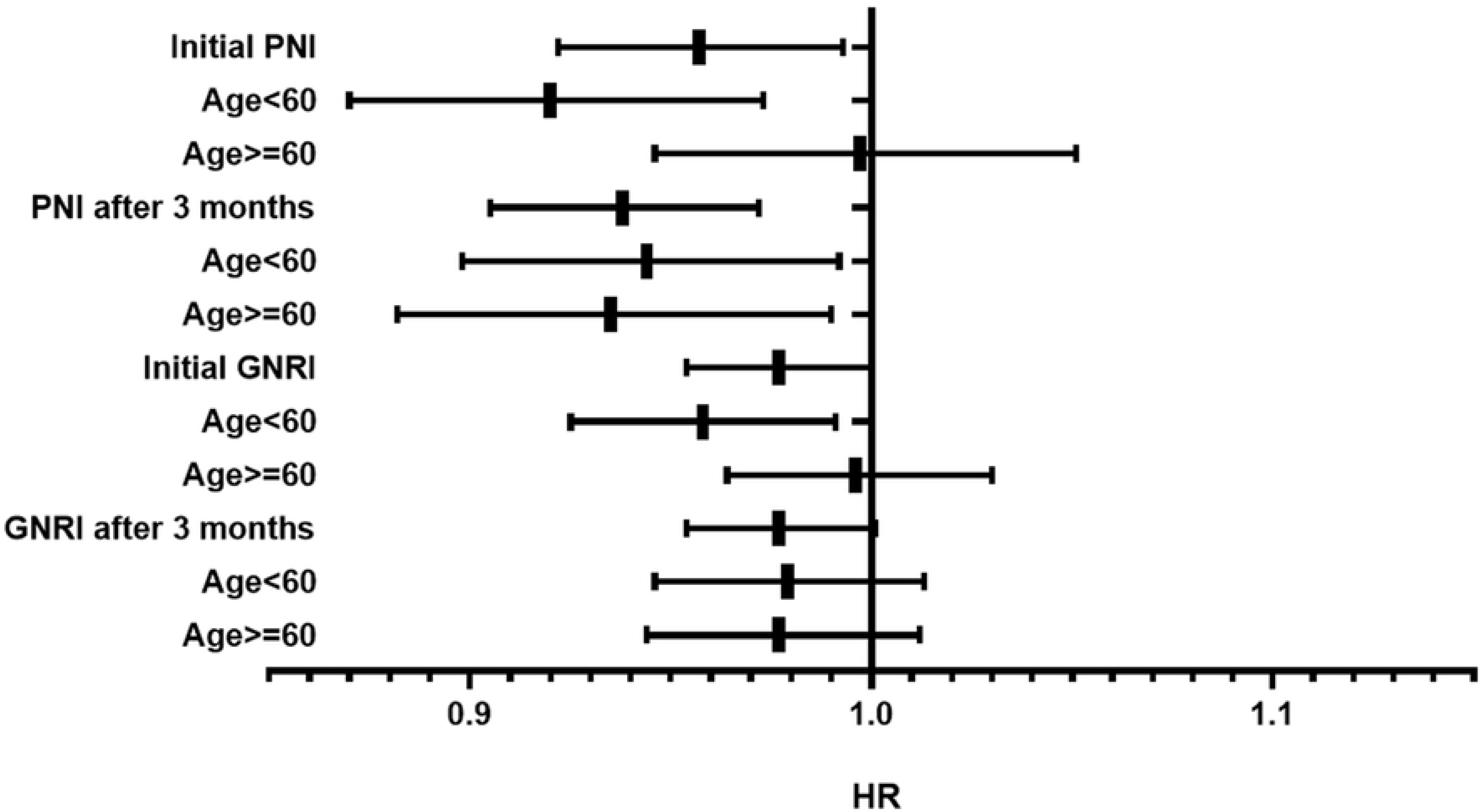
Impact of age on the predictive value of PNI and GNRI in patients undergoing hemodialysis. GNRI: geriatric nutritional risk index; PNI: prognostic nutritional index.

### The extent of the decrease in PNI before the patient’s death was more pronounced than that of GNRI

Among the 66 patients who died during the follow-up period, 63 patients underwent laboratory tests before death. The latest test was conducted from 2 months before death to 1 day before death. We collected the corresponding data and calculated the last PNI and GNRI before death. Among all 63 patients, the PNI decreased before death in 38 patients (60.31%), while the GNRI decreased in 33 patients (52.38%) before death. The former was slightly higher than the latter, but there was no statistically significant difference (p = 0.369). As shown in Fig 5, compared with the average PNI and GNRI at 3 months after the initiation of dialysis, both the average PNI and GNRI calculated before death indicated a decreasing trend. The decrease in average PNI was more pronounced and statistically significant (38.64 ± 7.63 vs. 41.27 ± 6.17, p = 0.03), while the decrease in average GNRI, although present, did not reach statistical significance compared with the average GNRI at 3 months after dialysis (90.93 ± 10.80 vs. 93.14 ± 8.76, p = 0.156). Despite the downward trends of average PNI and GNRI before death in the 63 patients, they were still higher than the baseline data.

**Fig 5.**
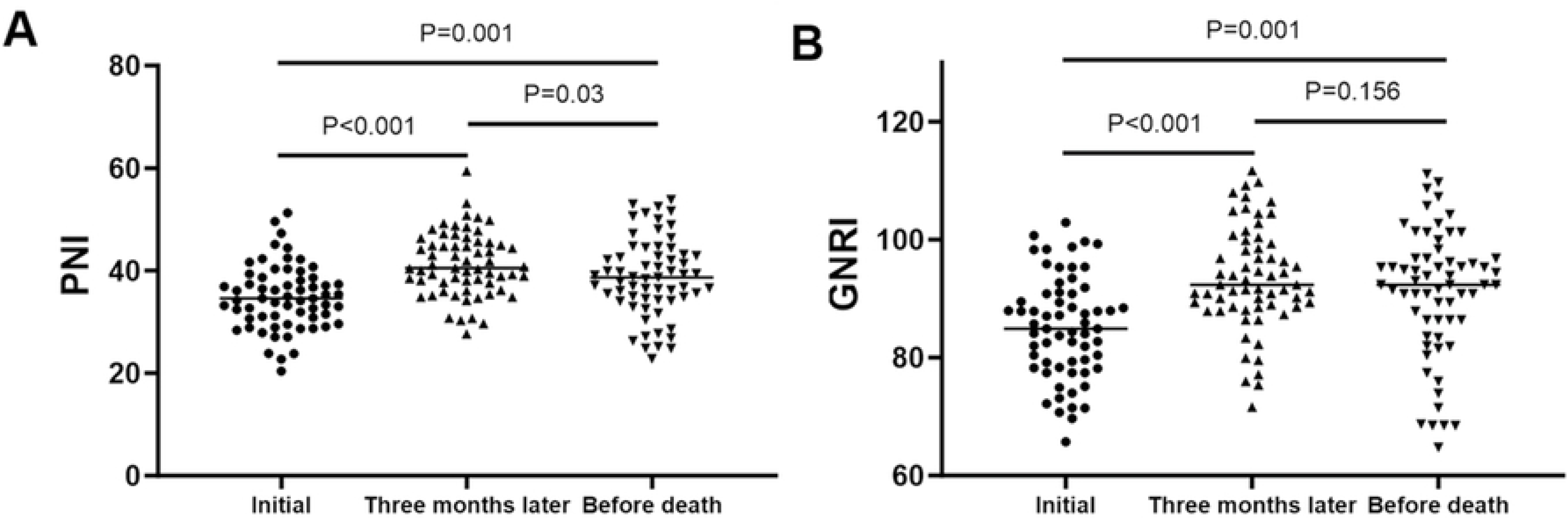
Comparison of PNI and GNRI values at dialysis initiation, three months after initiation, and before death in deceased patients.

## Discussion

MHD for patients with ESRD is a long-term treatment process, influenced by numerous factors that may affect prognosis [24]. Traditionally, prognostic predictions have required comprehensive analyses of various factors during MHD, including age, underlying health conditions, residual renal function, adequacy of dialysis, fluctuations in blood pressure, weight changes during dialysis sessions, and other relevant indices [25]. Consequently, identifying a simple indicator for early prediction holds considerable significance for MHD. Nutritional status serves as a critical indicator influencing the prognosis of many diseases [26]. In recent years, the GNRI has been utilized in some studies to evaluate the prognosis of MHD. However, it is important to note that GNRI was originally designed as an assessment tool specifically for elderly populations. Furthermore, the calculation formula for GNRI solely evaluates nutritional status without incorporating data related to immune function. ESRD patients typically experience immunocompromise which can lead to severe complications such as infections. PNI is an index that integrates the assessment of nutritional and immune status [27]. In recent years, it has been observed that PNI possesses considerable predictive value for various serious chronic diseases. A large-scale study conducted in the United States in 2023 highlighted the importance of PNI in forecasting outcomes for MHD [28]. Moreover, it was noted that PNI’s predictive capability surpassed that of absolute lymphocyte counts and serum albumin levels; however, this study did not address whether hemodialysis treatment itself influences PNI or explore how dynamic changes in PNI post-dialysis relate to patient prognosis.

In this study, we found that both the initial PNI level and the initial GNRI level are associated with patient prognosis, and the presence of diabetes significantly influences both indices. The proportion of diabetic patients in the low initial PNI group was markedly higher than that in the high initial PNI group. Similarly, a significantly greater proportion of diabetic patients was observed in the low initial GNRI group compared to the high initial GNRI group. We found that both PNI and GNRI at the initiation of dialysis were associated with long-term prognosis. However, since we excluded patients who had undergone dialysis for less than three months or lacked PNI and GNRI data at 3 months, we also examined whether these two indices were associated with early mortality. In fact, given the low rate of early mortality in our dialysis center, we did not find an association between these two indices and death within the first 3 months of dialysis. (Supplementary Fig 1).

Interestingly, although PNI is calculated based on blood lymphocyte counts, there were no significant differences in peripheral white blood cell or neutrophil counts between patients in the low and high initial PNI groups. Conversely, while GNRI calculation does not rely on immune cell counts, peripheral white blood cell and neutrophil counts were significantly elevated in patients within the low initial GNRI group compared to those with high levels. It is widely accepted that lymphocyte count serves as an indicator of immune function, whereas neutrophil count reflects inflammatory status. Therefore, a higher neutrophil count among patients with low initial GNRI may suggest an increased level of inflammation. Nutritional status is related to the prognosis of many diseases, while the mechanism by which immune function affects the prognosis of chronic diseases is more complex. The advantage of PNI over GNRI lies in the inclusion of the absolute value of lymphocytes, an indicator reflecting immune function. Kuwae N et al [29] examined the associations of the baseline white blood cell count and lymphocyte percentage with 12-month mortality and three measures of hospitalization in a cohort of 1655 MHD patients. They found a high WBC and a low lymphocyte percentage are associated with significant increase in mortality and hospitalization in patients. It is worth noting that the percentage of lymphocytes is not only related to prognosis but also to the “malnutrition-inflammation score”. Therefore, combining nutritional indicators with immune function to predict the prognosis of diseases seems more reasonable than using nutritional indicators alone.

MHD not only extends the lifespan of patients with uremia but also enhances their nutritional status and immune function [30]. Previous studies in non-dialysis patients have commonly used a PNI value below 38 as the cutoff for defining malnutrition [31, 32]. In our study, ROC analysis showed that when the PNI after three months of dialysis was set at 40.8, the Youden index for predicting mortality reached its maximum, indicating that nutritional status in hemodialysis patients must reach a relatively high level to achieve improved prognosis. We also observed that both the initial PNI and the PNI measured after three months of dialysis were associated with patient prognosis. However, the correlation between the PNI after three months of dialysis and prognosis was found to be more obvious than that between the initial PNI and prognosis. Additionally, while the initial GNRI level showed a relationship with prognosis, no significant correlation was identified between GNRI after three months of dialysis and prognosis. In multivariate Cox analysis, only the PNI assessed after three months of dialysis demonstrated a significant association with prognosis; conversely, neither initial PNI nor either measure of GNRI correlated significantly with prognostic outcomes thereafter.

Our findings indicate that following three months of hemodialysis, there was an increase in both PNI and GNRI levels among most patients: 79.5% exhibited increases in both indices, 13% experienced decreases in both measures, while 7.5% displayed inconsistent trends regarding changes in their PNI and GNRI values. The GNRI was originally developed for elderly patients; therefore, we conducted a comparative analysis of the predictive value of the PNI and GNRI regarding patient prognosis across different age groups. Surprisingly, our findings revealed that the initial GNRI demonstrated predictive value solely for patients under 60 years of age. The result of the poor predictive effect of GNRI in the elderly should be interpreted with caution. GNRI may be influenced by fluid status, dry weight estimation, and body composition changes, which could affect its prognostic performance in our cohort. It is worth noting that a study published in 2024, which included 863 hemodialysis patients, suggested that GNRI had higher predictive value for all-cause mortality than PNI [23]. We believe that differences in cohort population characteristics may partly account for the discrepant findings. In comparison, our cohort had a lower median age (55.0 vs. 60.0) and a higher proportion of patients with diabetes (39% vs. 19.7%). Therefore, further meta-analyses may be needed to validate these results.

There has been a scarcity of relevant studies regarding the alterations in PNI and GNRI before the death of dialysis patients. Therefore, we calculated the PNI and GNRI of these deceased patients before their death. We found that not all patients had a decrease in PNI and GNRI before death, but the overall average level was indeed lower than that three months after the start of dialysis. Compared with GNRI, the decline in PNI was more significant. Therefore, for hemodialysis patients, PNI may indeed be a more sensitive indicator than GNRI. However, given the substantial time span across which the pre-death data were collected, this finding should be categorized as exploratory, and further validation is warranted in future studies.

There are several limitations to this study that warrant clarification. First and most important, this study included only patients who survived and underwent reassessment at three months after dialysis initiation. Patients who died within three months of dialysis initiation or for whom PNI and GNRI data were unavailable at the three-month dialysis time point were excluded from the study. This may introduce potential survivor bias and influence the results. Second, the research was conducted at a single center and was a retrospective study, and there were no uniform or stringent protocols governing the hemodialysis regimen and medications administered to all patients. This may have compromised the generalizability of the study findings to other patient cohorts. Consequently, other factors influencing prognosis cannot be excluded. Third, due to the small sample size and irregular laboratory testing after more than three months of dialysis for some patients, we did not analyze the impact of PNI on prognosis following prolonged hemodialysis.

In summary, we found that both the initial PNI and the initial GNRI can serve as predictors for the long-term prognosis of patients undergoing MHD. After three months of hemodialysis, both PNI and GNRI showed an increase in most patients. However, only the PNI measured after three months of hemodialysis demonstrated a significant association with mortality. The relationship between the PNI at this time point and prognosis is more obvious than that observed with the initial PNI. Among the four indices evaluated, only the PNI following three months of hemodialysis is deemed suitable for elderly patients. The PNI after three months of MHD is a statistically significant but moderate predictor of long-term outcomes.

## Data Availability

All relevant data are within the manuscript and its Supporting Information files.

## Acknowledgments

We would like to express our sincere gratitude to everyone who supported this study, including the patients, doctors, and nursing team from the Hemodialysis Center of Tianjin Medical University General Hospital.

## Author contributions

Conceptualization: Junya Jia. Data curation: Lu Bai, Feifei Zhao.

Formal analysis: Liu Yang, Qing Zhang, Mingfan Lv. Funding acquisition: Pengcheng Xu, Junya Jia.

Investigation: Pengcheng Xu. Methodology: Haifei Xie, Fenxia Wang.

Resources: Yang Xue, Xiaoluan Liu. Supervision: Xinxin Zhang, Junya Jia. Validation: Shan Gao.

Visualization: Xinxin Zhang.

Writing – original draft: Shenghui Fu, Haomiao Zhang, Pengcheng Xu.

Writing – review & editing: Shenghui Fu, Haomiao Zhang, Pengcheng Xu, Junya Jia.

## Abbreviations

AUC: Area under the curve
BMI: Body mass index
CI: Confidence interval
CRP: C-reactive protein
eGFR: Estimated Glomerular filtration rate
ESRD: End-stage renal disease
GNRI: Geriatric Nutritional Risk Index
HDL: High-density lipoprotein
LDL: Low-density lipoprotein
MDRD: Modification of Diet in Renal Diseases
MHD: Maintenance hemodialysis
PNI: Prognostic Nutritional Index
ROC: Receiver operating characteristic

